# The 100 most-cited articles in soccer injuries: a bibliometric study

**DOI:** 10.1101/2023.04.27.23289221

**Authors:** Amr Chaabeni, Amine Kalai, Meryam Slama, Zohra Ben Salah Frih, Anis Jellad

## Abstract

Bibliometric analysis is an increasingly popular method for assessing aspects of a specific field of research and identifying its emerging areas of interest. Although it has been increasingly used in sports medicine research to assess injuries, few studies have focused on soccer. The purpose of our study was to identify the 100 most-cited publications related to soccer injuries and conduct a thorough bibliometric mapping analysis to understand the research trends. Using the Clarivate Analytics Web of Science Database and Bibliometrix R-package software, we identified 100 articles with 26,046 total citations (range = 146-945) published between 1990 and 2017. Most were published in the *British Journal of Sports Medicine* (32 articles) and *American Journal of Sports Medicine* (31 articles), and most corresponding authors originated from Sweden. About half of the articles were descriptive or epidemiology studies using level-2 evidence. The main research topics were epidemiology (25 articles) and prevention (24 articles). Most focused on male and adult elite soccer players.

## Introduction

Soccer is the most widely played sport in the world, with more than 265 million participants [1,2]. However, its practice is associated with a high rate of injuries, with an overall incidence of 8.1 injuries per 1,000 hours of practice [3]. It is estimated that professional soccer teams experience an average of 2 injuries per player in a single season [4]. As a result, professional teams and athletes may face significant challenges related to practice time loss and financial burdens [5]. This may explain many authors’ growing interest in the study of soccer injuries.

During the past decade, a growing number of bibliometric studies about sports have been published. Bibliometric studies are defined as a statistical assessment of published research aiming to quantify the impact and analyze the trends of publications, especially those with a high number of citations [6–8]. For research focused on soccer, bibliometric studies have focused on general topics, such as medicine and science in football [9,10]. To the best of our knowledge, no previous bibliometric studies have dealt with soccer injuries.

The purpose of our research was to identify the 100 most-cited publications related to soccer injuries and to carry out a thorough mapping analysis.

## Methods

We researched the Web of Science (WOS) database using the Clarivate Analytics tool in November 2022.

The search terms used were as follows:

*Topic Sentence (soccer OR football) NOT (American football OR Australian football OR rugby OR hockey OR cricket)*.

There were no restrictions related to the type of study, the language, or publication date.

Using the specified search terms, we found 44,527 articles on the WOS database. Articles were sorted by the number of citations. Three authors (AJ, AK and AC) assessed the first 300 articles; the reviewers individually verified the title and abstract of each article and kept only those dealing with injuries and traumatology that were related to soccer (football) athletes (articles comprising multiple sports were excluded). The analytical study was limited to the 100 most-cited articles.

### Data analysis

Bibliometrix R-package software was used for data analysis. The resulting 100 most-cited articles were analyzed to obtain the following variables: journal title, authors’ name, authors’ country of origin, year of publication, number of citations, citations per year, article type (original research, review article, descriptive/epidemiology article, case study, short communication, letter to the editor, editorial) [11] and level of evidence according to the standards set by the *Journal of Bone and Joint Surgery* [12,13]. Authors’ keywords were extracted to identify the trends.

Each article was assigned to topics based on the research question the authors attempted to address, such as epidemiology, anatomy and biomechanics, prevention, rehabilitation, classification and scoring, imaging, injury mechanism, surgical technique and outcome, and neuropsychology. Additionally, demographic information about the athletes, such as gender (male, female, or both), competition level (elite, non-elite, or both), and age category (youth under 18, adults over 19) were obtained from the original research articles. For the competition level, professional players or those playing in the first division were classified as “elite”, while amateur, college, or high-school players were classified as “non-elite” [9].

## Results

The 100 most-cited articles are listed in appendix 1 with their rank, number of citations and citations per year. These articles had a total of 26,046 citations, with an average of 260.5 citations per document (range: 146-945). The 100 most-cited articles were published in 21 journals. The two journals that published the most articles were the *British Journal of Sports Medicine* (32 articles, 8,425 citations) and the *American Journal of Sports Medicine* (31 articles, 8,119 citations) (table1).

The number of authors who contributed to the 100 most-cited articles was 279. The top 5 published authors were Hagglund, M. (20 articles); Ekstrand, J. (19 articles); Walden, M. (16 articles); Dvorak, J. (11 articles); and Junge, A. (10 articles). Authors who published 5 or more articles are presented in table 2. The top ten corresponding authors’ countries of origin are mentioned in figure 1 along with intra- and inter-countries collaboration. These collaborations occurred especially among European countries and other countries such as the United States, Canada, and Australia (figure 2).

**Table 1:** Distribution of journals per citations and articles

| **Journals** | **Number of**<br>**articles** | **Total**<br>**Citations** | **Publishing**<br>**Year start** |
| --- | --- | --- | --- |
| British Journal Of Sports Medicine | 32 | 8425 | 2001 |
| American Journal Of Sports Medicine | 31 | 8119 | 1991 |
| Scandinavian Journal Of Medicine \& Science In Sports | 9 | 2175 | 1996 |
| Knee Surgery Sports Traumatology Arthroscopy | 6 | 1471 | 2000 |
| Journal Of Athletic Training | 3 | 610 | 2007 |
| Sports Medicine | 3 | 585 | 1994 |
| Jama-Journal Of The American Medical Association | 2 | 433 | 1999 |
| Acta Orthopaedica Scandinavica | 1 | 162 | 1995 |
| Annals Of The Rheumatic Diseases | 1 | 557 | 2004 |
| Arthritis And Rheumatism | 1 | 945 | 2004 |
| Bmj-British Medical Journal | 1 | 228 | 2012 |
| British Medical Journal | 1 | 445 | 2008 |
| Clinical Journal Of Sport Medicine | 1 | 246 | 2006 |
| Foot & Ankle | 1 | 184 | 1990 |
| Journal Of Orthopaedic & Sports Physical Therapy | 1 | 244 | 2010 |
| Journal Of Pediatric Orthopaedics | 1 | 189 | 2004 |
| Journal Of Science And Medicine In Sport | 1 | 159 | 2010 |
| Lancet | 1 | 273 | 1999 |
| Medicine And Science In Sports And Exercise | 1 | 154 | 2003 |
| Neurology | 1 | 216 | 1998 |
| Radiology | 1 | 226 | 2013 |

**Table 2:** List of authors with 5 or more articles.

| **Authors** | **Number of**<br>**articles** | **Total**<br>**Citations** | **Publishing Year**<br>**start** |
| --- | --- | --- | --- |
| HAGGLUND M | 20 | 6208 | 2005 |
| EKSTRAND J | 19 | 5976 | 1990 |
| WALDEN M | 16 | 4764 | 2005 |
| DVORAK J | 11 | 2118 | 2000 |
| JUNGE A | 10 | 2520 | 2000 |
| ANDERSEN TE | 6 | 2271 | 2006 |
| BAHR R | 5 | 1729 | 2004 |
| HOLME I | 5 | 1742 | 2004 |

**Figure 1:**
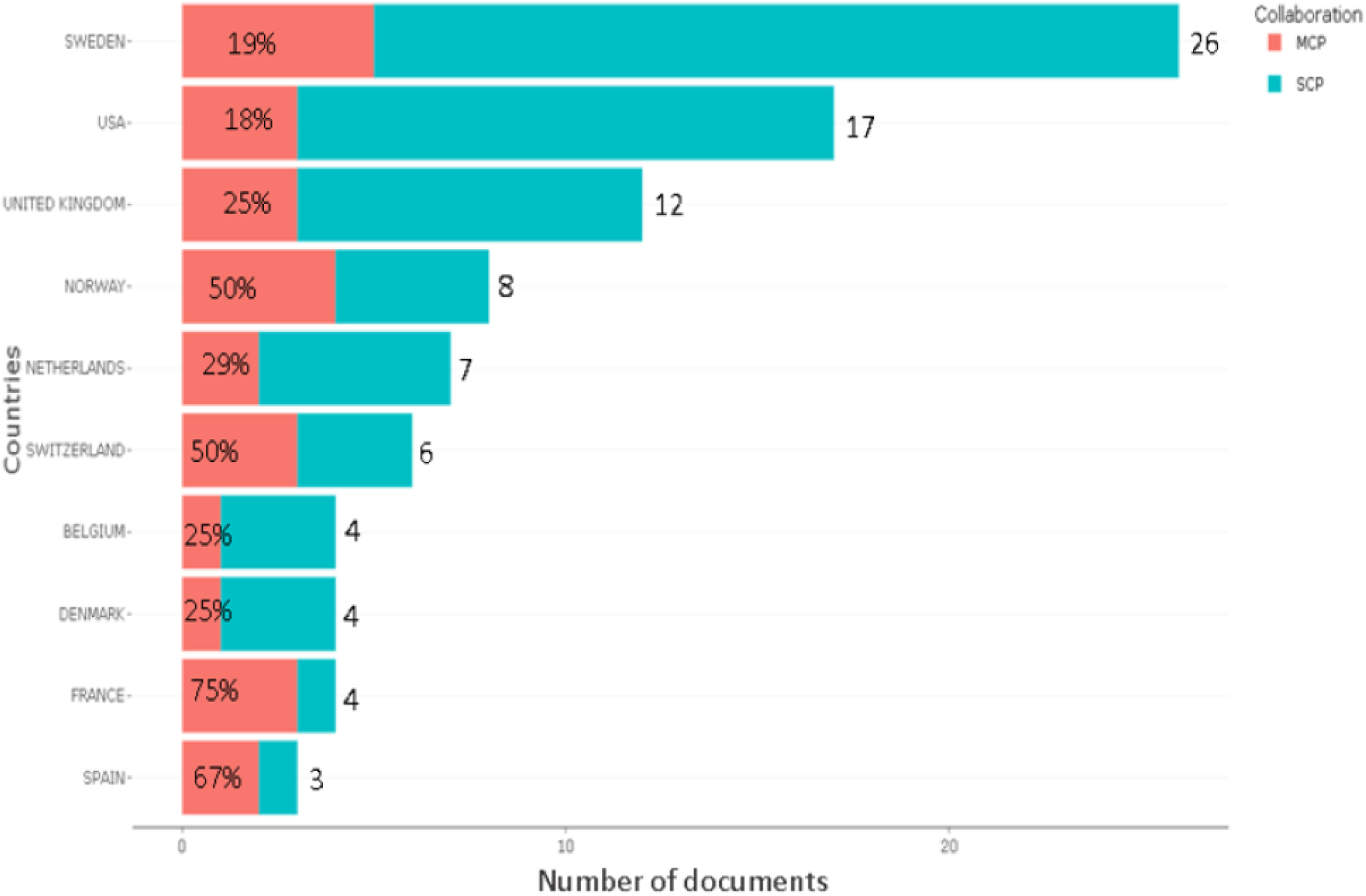
Number of articles per corresponding authors’ country of origin with intra-country (SCP) and inter-country (MCP) collaboration rates.

**Figure 2:**
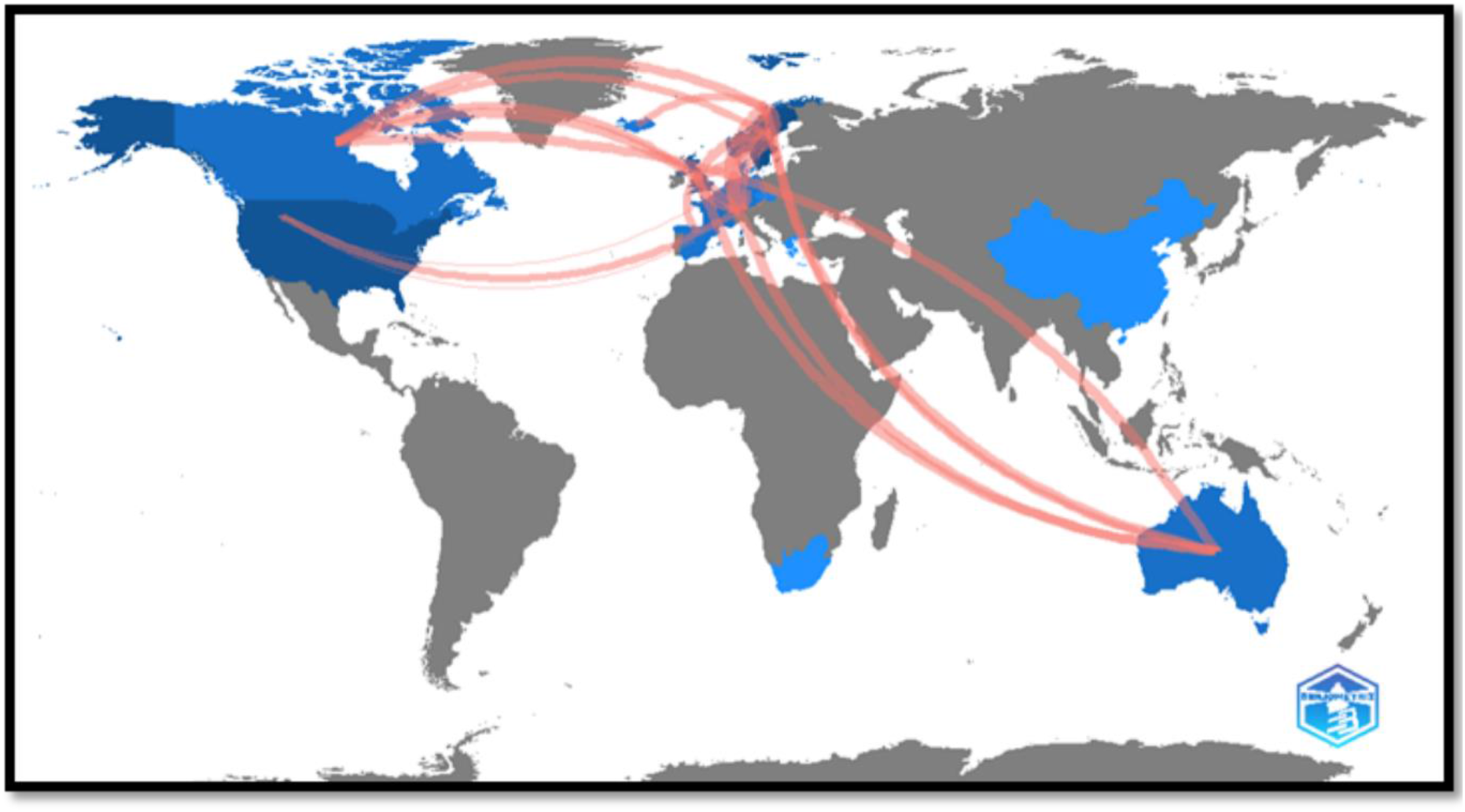
Collaboration inter-countries worldmap

The 100 most-cited articles were published between 1990 and 2017, with an average of 3.57 publications per year (figure 3). The mean number of citations per year per article was 17.65 (range: 5.75-39; figure 4). The most recent article was published in April 2017 and was the only meta-analysis in the list, with a total 146 citations and 24.33 citations per year. Of the 100 most-cited articles, 45 articles were descriptive or epidemiological, and 36 were original research. The level of evidence was one in 20 articles and two in 50 articles. The most common research topics are epidemiology in 25 articles, prevention in 24 articles, and injury risk factors in 20 articles. Most of the 100 most-cited articles were focused on male and adult elite soccer players (table 3).

**Table 3:** Descriptive data of the 100 Most-cited articles relating to soccer injuries.

| Variable | No. of articles |
| --- | --- |
| Article type |  |
| Descriptive/epidemiology | 45 |
| Original research | 36 |
| Review article | 16 |
| Case study | 2 |
| Letter to the editor | 1 |
| Level of evidence |  |
| 1 | 20 |
| 2 | 50 |
| 3 | 26 |
| 4 | 3 |
| 5 | 1 |
| Topic |  |
| Epidemiology | 25 |
| Prevention | 24 |
| Injury risk factors | 20 |
| Anatomy and biomechanics | 11 |
| Classification and scoring | 7 |
| Injury mechanism | 4 |
| Rehabilitation | 3 |
| Imaging | 2 |
| Surgical technique and outcome | 2 |
| Neuropsychology | 1 |
| Gender |  |
| Male | 53 |
| Female | 13 |
| Both | 34 |
| Age |  |
| Youth | 16 |
| Adults | 57 |
| Both | 27 |
| Competition level |  |
| Elite | 51 |
| Non-elite | 17 |
| Both | 32 |

**Figure 3:**
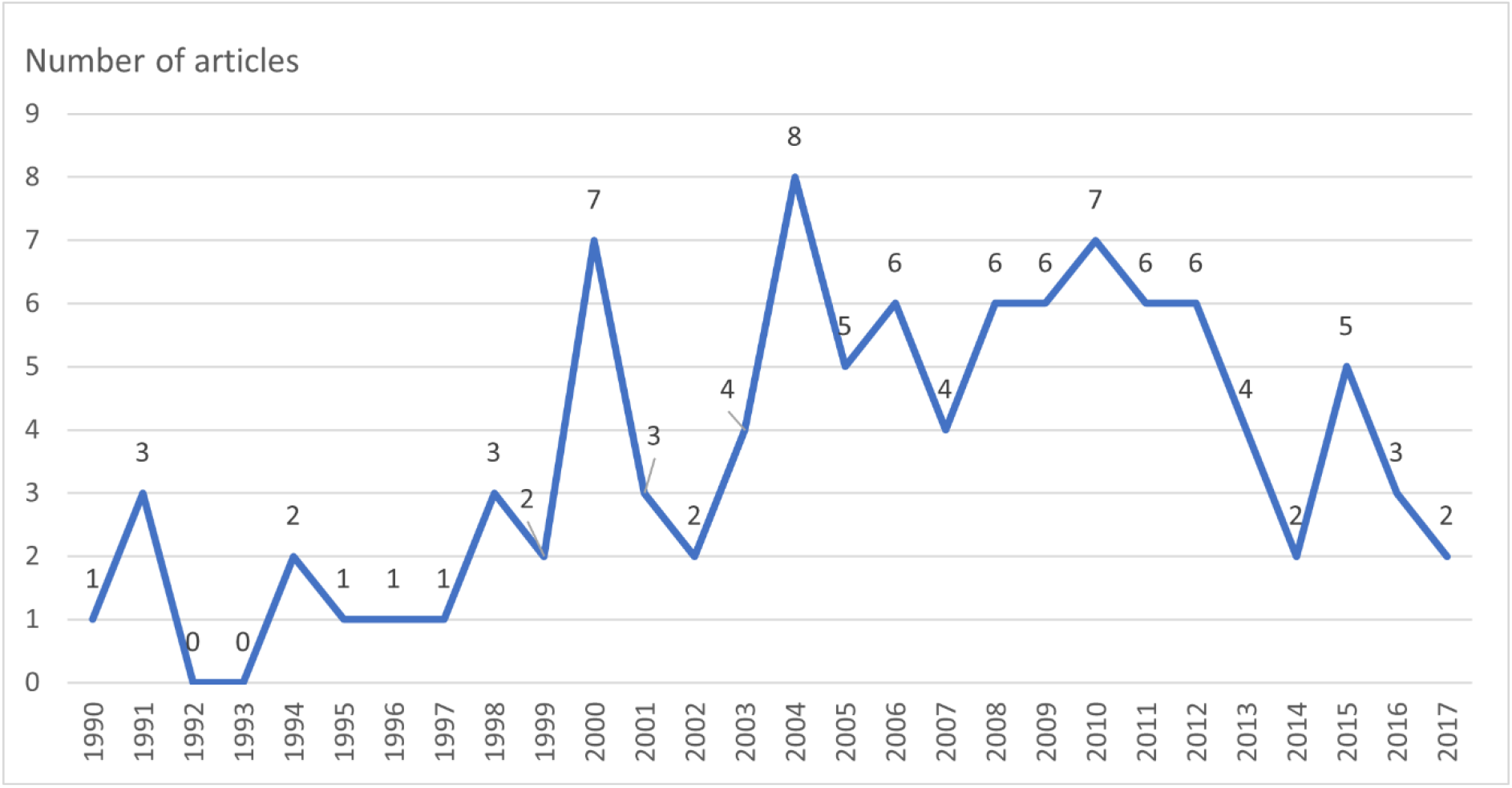
Annual production of the selected articles

**Figure 4:**
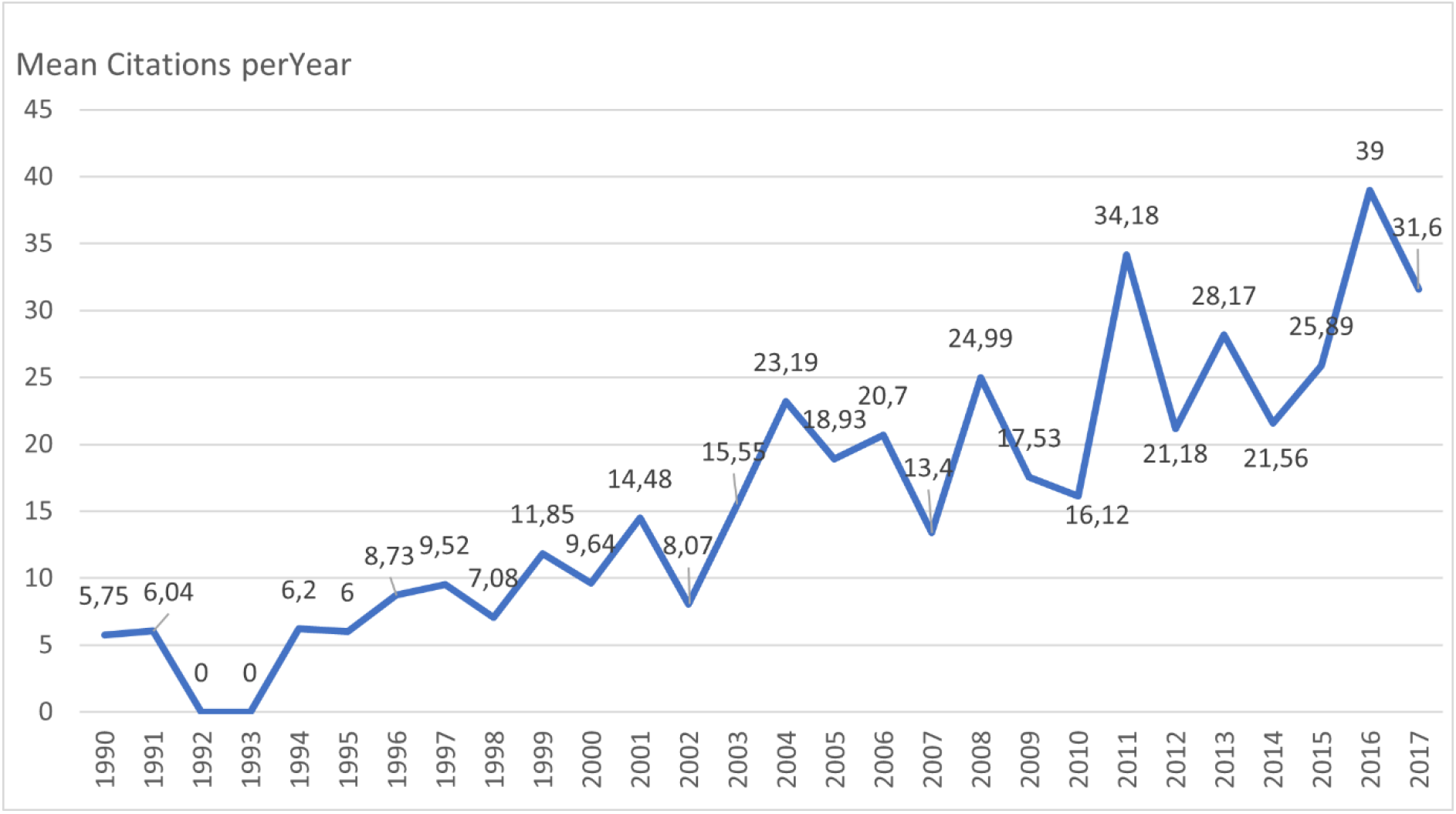
Mean number of citations per article per year.

## Discussion

This bibliometric study aimed to identify the 100 most-cited articles related to soccer injuries and determine their characteristics to understand the research and publishing trends in this area.

We found that the 100 most-cited articles were published between 1990 and 2017, mainly in the *British Journal of Sports Medicine* and the *American Journal of Sports Medicine*. In fact, the *American Journal of Sports Medicine* has shown a noticeable increase in the number of articles and citations in the past decades [14]. Furthermore, authors from high-ranking institutions who produce a high number of publications frequently publish in these journals. Thus, this may explain their high rate of citations [15,16]. Among the 5 most-cited authors, 3 (Hagglund, M., Ekstrand, J., and Walden, M.) are from Sweden and affiliated with the Linkoping University, and 2 (Dvorak, J. and Junge, A.) are affiliated with the FIFA Medical Assessment and Research Centre in Zurich, Switzerland. These 5 most-cited authors each have more than 200 articles published in the field of sports sciences. Moreover, the 3 most-cited authors (Hagglund, M., Ekstrand, J., and Walden, M.) were involved in writing the two articles with the highest number of citations per year (60.75 and 55.33) [4,17].

Descriptive and epidemiological studies were the most common among the 100 most-cited articles, and the main topics of the selected articles were epidemiology, prevention, and injury risk factors [18].

The major challenge for research in sports, especially in soccer, remains injury prevention [19,20]. Implementing prevention programs requires descriptive and epidemiological studies [21]. This may explain most authors’ interest in performing this type of study.

Despite the recent increase in the number of studies focusing on female players during the past two decades, fewer studies have focused on female players than male players [1]. Globally, elite athletes are studied more than recreational athletes. Their performance and financial imperatives may be the main explanation for these discrepancy [5,22].

## Conclusion

The 100 most-cited articles related to soccer injuries were mainly published in the *British Journal of Sports Medicine* and the *American Journal of Sports Medicine*. Most of these articles were descriptive and focused on epidemiology, risk factors, and prevention. Male players at the professional level were widely studied.

## Data Availability

All data produced in the present work are contained in the manuscript

