## Appendix 1 for "The 100 most-cited articles in soccer injuries: a bibliometric study"

| **Rank** | **Paper** | **No. of citations** | **No. of citations per year** |
| --- | --- | --- | --- |
| 1 | Lohmander LS, Ostenberg A, Englund M, Roos H. High prevalence of knee osteoarthritis, pain, and functional limitations in female soccer players twelve years after anterior cruciate ligament injury. Arthritis Rheum. 2004 Oct;50(10):3145-52. | 945 | 49,74 |
| 2 | Fuller CW, Ekstrand J, Junge A, Andersen TE, Bahr R, Dvorak J, Hägglund M, McCrory P, Meeuwisse WH. Consensus statement on injury definitions and data collection procedures in studies of football (soccer) injuries. Br J Sports Med. 2006 Mar;40(3):193-201. | 794 | 46,71 |
| 3 | Ekstrand J, Hägglund M, Waldén M. Injury incidence and injury patterns in professional football: the UEFA injury study. Br J Sports Med. 2011 Jun;45(7):553-8. | 729 | 60,75 |
| 4 | Ekstrand J, Hägglund M, Waldén M. Epidemiology of muscle injuries in professional football (soccer). Am J Sports Med. 2011 Jun;39(6):1226-32. | 664 | 55,33 |
| 5 | Mandelbaum BR, Silvers HJ, Watanabe DS, Knarr JF, Thomas SD, Griffin LY, Kirkendall DT, Garrett W Jr. Effectiveness of a neuromuscular and proprioceptive training program in preventing anterior cruciate ligament injuries in female athletes: 2-year follow-up. Am J Sports Med. 2005 Jul;33(7):1003-10. | 664 | 36,89 |
| 6 | Croisier JL, Ganteaume S, Binet J, Genty M, Ferret JM. Strength imbalances and prevention of hamstring injury in professional soccer players: a prospective study. Am J Sports Med. 2008 Aug;36(8):1469-75. | 559 | 37,27 |
| 7 | von Porat A, Roos EM, Roos H. High prevalence of osteoarthritis 14 years after an anterior cruciate ligament tear in male soccer players: a study of radiographic and patient relevant outcomes. Ann Rheum Dis. 2004 Mar;63(3):269-73. | 557 | 29,32 |
| 8 | Woods C, Hawkins RD, Maltby S, Hulse M, Thomas A, Hodson A; Football Association Medical Research Programme. The Football Association Medical Research Programme: an audit of injuries in professional football--analysis of hamstring injuries. Br J Sports Med. 2004 Feb;38(1):36-41. | 555 | 29,21 |
| 9 | Arnason A, Sigurdsson SB, Gudmundsson A, Holme I, Engebretsen L, Bahr R. Risk factors for injuries in football. Am J Sports Med. 2004 Jan-Feb;32(1 Suppl):5S-16S. | 536 | 28,21 |
| 10 | Alentorn-Geli E, Myer GD, Silvers HJ, Samitier G, Romero D, Lázaro-Haro C, Cugat R. Prevention of non-contact anterior cruciate ligament injuries in soccer players. Part 1: Mechanisms of injury and underlying risk factors. Knee Surg Sports Traumatol Arthrosc. 2009 Jul;17(7):705-29. | 487 | 34,79 |
| 11 | Hawkins RD, Hulse MA, Wilkinson C, Hodson A, Gibson M. The association football medical research programme: an audit of injuries in professional football. Br J Sports Med. 2001 Feb;35(1):43-7. | 472 | 21,45 |
| 12 | Askling C, Karlsson J, Thorstensson A. Hamstring injury occurrence in elite soccer players after preseason strength training with eccentric overload. Scand J Med Sci Sports. 2003 Aug;13(4):244-50. | 469 | 23,45 |
| 13 | Soligard T, Myklebust G, Steffen K, Holme I, Silvers H, Bizzini M, Junge A, Dvorak J, Bahr R, Andersen TE. Comprehensive warm-up programme to prevent injuries in young female footballers: cluster randomised controlled trial. BMJ. 2008 Dec 9;337:a2469. | 445 | 29,67 |
| 14 | Witvrouw E, Danneels L, Asselman P, D'Have T, Cambier D. Muscle flexibility as a risk factor for developing muscle injuries in male professional soccer players. A prospective study. Am J Sports Med. 2003 Jan-Feb;31(1):41-6. | 399 | 19,95 |
| 15 | Hägglund M, Waldén M, Ekstrand J. Previous injury as a risk factor for injury in elite football: a prospective study over two consecutive seasons. Br J Sports Med. 2006 Sep;40(9):767-72. | 371 | 21,82 |
| 16 | Hägglund M, Waldén M, Magnusson H, Kristenson K, Bengtsson H, Ekstrand J. Injuries affect team performance negatively in professional football: an 11-year follow-up of the UEFA Champions League injury study. Br J Sports Med. 2013 Aug;47(12):738-42. | 370 | 37,00 |
| 17 | Arnason A, Andersen TE, Holme I, Engebretsen L, Bahr R. Prevention of hamstring strains in elite soccer: an intervention study. Scand J Med Sci Sports. 2008 Feb;18(1):40-8. | 349 | 23,27 |
| 18 | Petersen J, Thorborg K, Nielsen MB, Budtz-Jørgensen E, Hölmich P. Preventive effect of eccentric training on acute hamstring injuries in men's soccer: a cluster-randomized controlled trial. Am J Sports Med. 2011 Nov;39(11):2296-303. | 344 | 28,67 |
| 19 | Hägglund M, Waldén M, Bahr R, Ekstrand J. Methods for epidemiological study of injuries to professional football players: developing the UEFA model. Br J Sports Med. 2005 Jun;39(6):340-6. | 341 | 18,94 |
| 20 | Gilchrist J, Mandelbaum BR, Melancon H, Ryan GW, Silvers HJ, Griffin LY, Watanabe DS, Dick RW, Dvorak J. A randomized controlled trial to prevent noncontact anterior cruciate ligament injury in female collegiate soccer players. Am J Sports Med. 2008 Aug;36(8):1476-83. | 339 | 22,60 |
| 21 | Ekstrand J, Waldén M, Hägglund M. Hamstring injuries have increased by 4% annually in men's professional football, since 2001: a 13-year longitudinal analysis of the UEFA Elite Club injury study. Br J Sports Med. 2016 Jun;50(12):731-7. | 330 | 47,14 |
| 22 | Heidt RS Jr, Sweeterman LM, Carlonas RL, Traub JA, Tekulve FX. Avoidance of soccer injuries with preseason conditioning. Am J Sports Med. 2000 Sep-Oct;28(5):659-62. | 285 | 12,39 |
| 23 | Hölmich P, Uhrskou P, Ulnits L, Kanstrup IL, Nielsen MB, Bjerg AM, Krogsgaard K. Effectiveness of active physical training as treatment for long-standing adductor-related groin pain in athletes: randomised trial. Lancet. 1999 Feb 6;353(9151):439-43. | 273 | 11,38 |
| 24 | Matser EJ, Kessels AG, Lezak MD, Jordan BD, Troost J. Neuropsychological impairment in amateur soccer players. JAMA. 1999 Sep 8;282(10):971-3. | 272 | 11,33 |
| 25 | Waldén M, Hägglund M, Ekstrand J. UEFA Champions League study: a prospective study of injuries in professional football during the 2001-2002 season. Br J Sports Med. 2005 Aug;39(8):542-6. | 267 | 14,83 |
| 26 | Ekstrand J, Healy JC, Waldén M, Lee JC, English B, Hägglund M. Hamstring muscle injuries in professional football: the correlation of MRI findings with return to play. Br J Sports Med. 2012 Feb;46(2):112-7. | 259 | 23,55 |
| 27 | Dupont G, Nedelec M, McCall A, McCormack D, Berthoin S, Wisløff U. Effect of 2 soccer matches in a week on physical performance and injury rate. Am J Sports Med. 2010 Sep;38(9):1752-8. | 257 | 19,77 |
| 28 | Yu B, Garrett WE. Mechanisms of non-contact ACL injuries. Br J Sports Med. 2007 Aug;41 Suppl 1(Suppl 1):i47-51. | 254 | 15,88 |
| 29 | Fuller CW, Ekstrand J, Junge A, Andersen TE, Bahr R, Dvorak J, Hägglund M, McCrory P, Meeuwisse WH. Consensus statement on injury definitions and data collection procedures in studies of football (soccer) injuries. Scand J Med Sci Sports. 2006 Apr;16(2):83-92. | 249 | 14,65 |
| 30 | Fuller CW, Ekstrand J, Junge A, Andersen TE, Bahr R, Dvorak J, Hägglund M, McCrory P, Meeuwisse WH. Consensus statement on injury definitions and data collection procedures in studies of football (soccer) injuries. Clin J Sport Med. 2006 Mar;16(2):97-106. | 246 | 14,47 |
| 31 | Heiderscheit BC, Sherry MA, Silder A, Chumanov ES, Thelen DG. Hamstring strain injuries: recommendations for diagnosis, rehabilitation, and injury prevention. J Orthop Sports Phys Ther. 2010 Feb;40(2):67-81. | 244 | 18,77 |
| 32 | Söderman K, Werner S, Pietilä T, Engström B, Alfredson H. Balance board training: prevention of traumatic injuries of the lower extremities in female soccer players? A prospective randomized intervention study. Knee Surg Sports Traumatol Arthrosc. 2000;8(6):356-63. | 239 | 10,39 |
| 33 | Nédélec M, McCall A, Carling C, Legall F, Berthoin S, Dupont G. Recovery in soccer: part I - post-match fatigue and time course of recovery. Sports Med. 2012 Dec 1;42(12):997-1015. | 238 | 21,64 |
| 34 | Bjordal JM, Arnły F, Hannestad B, Strand T. Epidemiology of anterior cruciate ligament injuries in soccer. Am J Sports Med. 1997 May-Jun;25(3):341-5. | 238 | 9,15 |
| 35 | Steffen K, Myklebust G, Olsen OE, Holme I, Bahr R. Preventing injuries in female youth football--a cluster-randomized controlled trial. Scand J Med Sci Sports. 2008 Oct;18(5):605-14. | 230 | 15,33 |
| 36 | Söderman K, Alfredson H, Pietilä T, Werner S. Risk factors for leg injuries in female soccer players: a prospective investigation during one out-door season. Knee Surg Sports Traumatol Arthrosc. 2001 Sep;9(5):313-21. | 230 | 10,45 |
| 37 | Waldén M, Atroshi I, Magnusson H, Wagner P, Hägglund M. Prevention of acute knee injuries in adolescent female football players: cluster randomised controlled trial. BMJ. 2012 May 3;344:e3042. | 228 | 20,73 |
| 38 | Peterson L, Junge A, Chomiak J, Graf-Baumann T, Dvorak J. Incidence of football injuries and complaints in different age groups and skill-level groups. Am J Sports Med. 2000;28(5 Suppl):S51-7. | 228 | 9,91 |
| 39 | Arnason A, Gudmundsson A, Dahl HA, Jóhannsson E. Soccer injuries in Iceland. Scand J Med Sci Sports. 1996 Feb;6(1):40-5. | 227 | 8,41 |
| 40 | Lipton ML, Kim N, Zimmerman ME, Kim M, Stewart WF, Branch CA, Lipton RB. Soccer heading is associated with white matter microstructural and cognitive abnormalities. Radiology. 2013 Sep;268(3):850-7. | 226 | 22,60 |
| 41 | Colvin AC, Mullen J, Lovell MR, West RV, Collins MW, Groh M. The role of concussion history and gender in recovery from soccer-related concussion. Am J Sports Med. 2009 Sep;37(9):1699-704. | 226 | 16,14 |
| 42 | Boytim MJ, Fischer DA, Neumann L. Syndesmotic ankle sprains. Am J Sports Med. 1991 May-Jun;19(3):294-8. | 225 | 7,03 |
| 43 | Matser JT, Kessels AG, Jordan BD, Lezak MD, Troost J. Chronic traumatic brain injury in professional soccer players. Neurology. 1998 Sep;51(3):791-6. | 216 | 8,64 |
| 44 | Brophy RH, Schmitz L, Wright RW, Dunn WR, Parker RD, Andrish JT, McCarty EC, Spindler KP. Return to play and future ACL injury risk after ACL reconstruction in soccer athletes from the Multicenter Orthopaedic Outcomes Network (MOON) group. Am J Sports Med. 2012 Nov;40(11):2517-22. | 215 | 19,55 |
| 45 | Timmins RG, Bourne MN, Shield AJ, Williams MD, Lorenzen C, Opar DA. Short biceps femoris fascicles and eccentric knee flexor weakness increase the risk of hamstring injury in elite football (soccer): a prospective cohort study. Br J Sports Med. 2016 Dec;50(24):1524-1535. | 213 | 30,43 |
| 46 | Steffen K, Emery CA, Romiti M, Kang J, Bizzini M, Dvorak J, Finch CF, Meeuwisse WH. High adherence to a neuromuscular injury prevention programme (FIFA 11+) improves functional balance and reduces injury risk in Canadian youth female football players: a cluster randomised trial. Br J Sports Med. 2013 Aug;47(12):794-802. | 212 | 21,20 |
| 47 | Agel J, Evans TA, Dick R, Putukian M, Marshall SW. Descriptive epidemiology of collegiate men's soccer injuries: National Collegiate Athletic Association Injury Surveillance System, 1988-1989 through 2002-2003. J Athl Train. 2007 Apr-Jun;42(2):270-7. | 210 | 13,13 |
| 48 | Price RJ, Hawkins RD, Hulse MA, Hodson A. The Football Association medical research programme: an audit of injuries in academy youth football. Br J Sports Med. 2004 Aug;38(4):466-71. | 210 | 11,05 |
| 49 | Drawer S, Fuller CW. Propensity for osteoarthritis and lower limb joint pain in retired professional soccer players. Br J Sports Med. 2001 Dec;35(6):402-8. | 210 | 9,55 |
| 50 | Padua DA, DiStefano LJ, Beutler AI, de la Motte SJ, DiStefano MJ, Marshall SW. The Landing Error Scoring System as a Screening Tool for an Anterior Cruciate Ligament Injury-Prevention Program in Elite-Youth Soccer Athletes. J Athl Train. 2015 Jun;50(6):589-95. | 207 | 25,88 |
| 51 | Hägglund M, Waldén M, Ekstrand J. Risk factors for lower extremity muscle injury in professional soccer: the UEFA Injury Study. Am J Sports Med. 2013 Feb;41(2):327-35. | 206 | 20,60 |
| 52 | van der Horst N, Smits DW, Petersen J, Goedhart EA, Backx FJ. The preventive effect of the nordic hamstring exercise on hamstring injuries in amateur soccer players: a randomized controlled trial. Am J Sports Med. 2015 Jun;43(6):1316-23. | 205 | 25,63 |
| 53 | Croisier JL. Factors associated with recurrent hamstring injuries. Sports Med. 2004;34(10):681-95. | 198 | 10,42 |
| 54 | Surve I, Schwellnus MP, Noakes T, Lombard C. A fivefold reduction in the incidence of recurrent ankle sprains in soccer players using the Sport-Stirrup orthosis. Am J Sports Med. 1994 Sep-Oct;22(5):601-6. | 198 | 6,83 |
| 55 | Dvorak J, Junge A. Football injuries and physical symptoms. A review of the literature. Am J Sports Med. 2000;28(5 Suppl):S3-9. | 196 | 8,52 |
| 56 | Dick R, Putukian M, Agel J, Evans TA, Marshall SW. Descriptive epidemiology of collegiate women's soccer injuries: National Collegiate Athletic Association Injury Surveillance System, 1988-1989 through 2002-2003. J Athl Train. 2007 Apr-Jun;42(2):278-85. | 193 | 12,06 |
| 57 | Ostenberg A, Roos H. Injury risk factors in female European football. A prospective study of 123 players during one season. Scand J Med Sci Sports. 2000 Oct;10(5):279-85. | 192 | 8,35 |
| 58 | Fousekis K, Tsepis E, Poulmedis P, Athanasopoulos S, Vagenas G. Intrinsic risk factors of non-contact quadriceps and hamstring strains in soccer: a prospective study of 100 professional players. Br J Sports Med. 2011 Jul;45(9):709-14. | 189 | 15,75 |
| 59 | Shea KG, Pfeiffer R, Wang JH, Curtin M, Apel PJ. Anterior cruciate ligament injury in pediatric and adolescent soccer players: an analysis of insurance data. J Pediatr Orthop. 2004 Nov-Dec;24(6):623-8 | 189 | 9,95 |
| 60 | Waldén M, Krosshaug T, Bjørneboe J, Andersen TE, Faul O, Hägglund M. Three distinct mechanisms predominate in non-contact anterior cruciate ligament injuries in male professional football players: a systematic video analysis of 39 cases. Br J Sports Med. 2015 Nov;49(22):1452-60 | 188 | 23,50 |
| 61 | Alentorn-Geli E, Myer GD, Silvers HJ, Samitier G, Romero D, Lázaro-Haro C, Cugat R. Prevention of non-contact anterior cruciate ligament injuries in soccer players. Part 2: a review of prevention programs aimed to modify risk factors and to reduce injury rates. Knee Surg Sports Traumatol Arthrosc. 2009 Aug;17(8):859-79. | 185 | 13,21 |
| 62 | Ekstrand J, Tropp H. The incidence of ankle sprains in soccer. Foot Ankle. 1990 Aug;11(1):41-4. | 184 | 5,58 |
| 63 | Soligard T, Nilstad A, Steffen K, Myklebust G, Holme I, Dvorak J, Bahr R, Andersen TE. Compliance with a comprehensive warm-up programme to prevent injuries in youth football. Br J Sports Med. 2010 Sep;44(11):787-93. | 182 | 14,00 |
| 64 | Chomiak J, Junge A, Peterson L, Dvorak J. Severe injuries in football players. Influencing factors. Am J Sports Med. 2000;28(5 Suppl):S58-68. | 182 | 7,91 |
| 65 | Engström B, Johansson C, Törnkvist H. Soccer injuries among elite female players. Am J Sports Med. 1991 Jul-Aug;19(4):372-5. | 180 | 5,63 |
| 66 | Agricola R, Heijboer MP, Ginai AZ, Roels P, Zadpoor AA, Verhaar JA, Weinans H, Waarsing JH. A cam deformity is gradually acquired during skeletal maturation in adolescent and young male soccer players: a prospective study with minimum 2-year follow-up. Am J Sports Med. 2014 Apr;42(4):798-806. | 178 | 19,78 |
| 67 | Engebretsen AH, Myklebust G, Holme I, Engebretsen L, Bahr R. Prevention of injuries among male soccer players: a prospective, randomized intervention study targeting players with previous injuries or reduced function. Am J Sports Med. 2008 Jun;36(6):1052-60. | 177 | 11,80 |
| 68 | Wong P, Hong Y. Soccer injury in the lower extremities. Br J Sports Med. 2005 Aug;39(8):473-82. | 176 | 9,78 |
| 69 | Emery CA, Meeuwisse WH. The effectiveness of a neuromuscular prevention strategy to reduce injuries in youth soccer: a cluster-randomised controlled trial. Br J Sports Med. 2010 Jun;44(8):555-62. | 175 | 13,46 |
| 70 | Waldén M, Hägglund M, Magnusson H, Ekstrand J. Anterior cruciate ligament injury in elite football: a prospective three-cohort study. Knee Surg Sports Traumatol Arthrosc. 2011 Jan;19(1):11-9. | 173 | 14,42 |
| 71 | Brophy R, Silvers HJ, Gonzales T, Mandelbaum BR. Gender influences: the role of leg dominance in ACL injury among soccer players. Br J Sports Med. 2010 Aug;44(10):694-7. | 172 | 13,23 |
| 72 | Le Gall F, Carling C, Reilly T, Vandewalle H, Church J, Rochcongar P. Incidence of injuries in elite French youth soccer players: a 10-season study. Am J Sports Med. 2006 Jun;34(6):928-38. | 172 | 10,12 |
| 73 | Woods C, Hawkins R, Hulse M, Hodson A. The Football Association Medical Research Programme: an audit of injuries in professional football-analysis of preseason injuries. Br J Sports Med. 2002 Dec;36(6):436-41; discussion 441. | 171 | 8,14 |
| 74 | Bowen L, Gross AS, Gimpel M, Li FX. Accumulated workloads and the acute:chronic workload ratio relate to injury risk in elite youth football players. Br J Sports Med. 2017 Mar;51(5):452-459. | 170 | 28,33 |
| 75 | Agricola R, Bessems JH, Ginai AZ, Heijboer MP, van der Heijden RA, Verhaar JA, Weinans H, Waarsing JH. The development of Cam-type deformity in adolescent and young male soccer players. Am J Sports Med. 2012 May;40(5):1099-106. | 170 | 15,45 |
| 76 | McCall A, Carling C, Nedelec M, Davison M, Le Gall F, Berthoin S, Dupont G. Risk factors, testing and preventative strategies for non-contact injuries in professional football: current perceptions and practices of 44 teams from various premier leagues. Br J Sports Med. 2014 Sep;48(18):1352-7. | 167 | 18,56 |
| 77 | Brink MS, Visscher C, Arends S, Zwerver J, Post WJ, Lemmink KA. Monitoring stress and recovery: new insights for the prevention of injuries and illnesses in elite youth soccer players. Br J Sports Med. 2010 Sep;44(11):809-15. | 165 | 12,69 |
| 78 | Hägglund M, Waldén M, Ekstrand J. Injuries among male and female elite football players. Scand J Med Sci Sports. 2009 Dec;19(6):819-27. | 165 | 11,79 |
| 79 | Junge A, Dvorak J. Influence of definition and data collection on the incidence of injuries in football. Am J Sports Med. 2000;28(5 Suppl):S40-6. | 163 | 7,09 |
| 80 | Roos H, Ornell M, Gärdsell P, Lohmander LS, Lindstrand A. Soccer after anterior cruciate ligament injury--an incompatible combination? A national survey of incidence and risk factors and a 7-year follow-up of 310 players. Acta Orthop Scand. 1995 Apr;66(2):107-12. | 162 | 5,79 |
| 81 | Faude O, Junge A, Kindermann W, Dvorak J. Injuries in female soccer players: a prospective study in the German national league. Am J Sports Med. 2005 Nov;33(11):1694-700. | 161 | 8,94 |
| 82 | Koerte IK, Ertl-Wagner B, Reiser M, Zafonte R, Shenton ME. White matter integrity in the brains of professional soccer players without a symptomatic concussion. JAMA. 2012 Nov 14;308(18):1859-61. | 161 | 14,64 |
| 83 | Woods C, Hawkins R, Hulse M, Hodson A. The Football Association Medical Research Programme: an audit of injuries in professional football: an analysis of ankle sprains. Br J Sports Med. 2003 Jun;37(3):233-8. | 160 | 8,00 |
| 84 | Waldén M, Hägglund M, Magnusson H, Ekstrand J. ACL injuries in men's professional football: a 15-year prospective study on time trends and return-to-play rates reveals only 65% of players still play at the top level 3 years after ACL rupture. Br J Sports Med. 2016 Jun;50(12):744-50. | 159 | 22,71 |
| 85 | Small K, McNaughton L, Greig M, Lovell R. The effects of multidirectional soccer-specific fatigue on markers of hamstring injury risk. J Sci Med Sport. 2010 Jan;13(1):120-5. | 159 | 12,23 |
| 86 | Waldén M, Hägglund M, Werner J, Ekstrand J. The epidemiology of anterior cruciate ligament injury in football (soccer): a review of the literature from a gender-related perspective. Knee Surg Sports Traumatol Arthrosc. 2011 Jan;19(1):3-10. | 157 | 13,08 |
| 87 | Werner J, Hägglund M, Waldén M, Ekstrand J. UEFA injury study: a prospective study of hip and groin injuries in professional football over seven consecutive seasons. Br J Sports Med. 2009 Dec;43(13):1036-40. | 157 | 11,21 |
| 88 | Tysvaer AT, Løchen EA. Soccer injuries to the brain. A neuropsychologic study of former soccer players. Am J Sports Med. 1991 Jan-Feb;19(1):56-60. | 157 | 4,91 |
| 89 | Ekstrand J, Timpka T, Hägglund M. Risk of injury in elite football played on artificial turf versus natural grass: a prospective two-cohort study. Br J Sports Med. 2006 Dec;40(12):975-80. | 155 | 9,12 |
| 90 | Bizzini M, Dvorak J. FIFA 11+: an effective programme to prevent football injuries in various player groups worldwide-a narrative review. Br J Sports Med. 2015 May;49(9):577-9. | 154 | 19,25 |
| 91 | Sánchez M, Azofra J, Anitua E, Andía I, Padilla S, Santisteban J, Mujika I. Plasma rich in growth factors to treat an articular cartilage avulsion: a case report. Med Sci Sports Exerc. 2003 Oct;35(10):1648-52. | 154 | 7,70 |
| 92 | Silvers-Granelli H, Mandelbaum B, Adeniji O, Insler S, Bizzini M, Pohlig R, Junge A, Snyder-Mackler L, Dvorak J. Efficacy of the FIFA 11+ Injury Prevention Program in the Collegiate Male Soccer Player. Am J Sports Med. 2015 Nov;43(11):2628-37. | 152 | 19,00 |
| 93 | Rahnama N, Reilly T, Lees A. Injury risk associated with playing actions during competitive soccer. Br J Sports Med. 2002 Oct;36(5):354-9. | 152 | 7,24 |
| 94 | Junge A, Dvorak J, Graf-Baumann T, Peterson L. Football injuries during FIFA tournaments and the Olympic Games, 1998-2001: development and implementation of an injury-reporting system. Am J Sports Med. 2004 Jan-Feb;32(1 Suppl):80S-9S. | 149 | 7,84 |
| 95 | Inklaar H. Soccer injuries. I: Incidence and severity. Sports Med. 1994 Jul;18(1):55-73. | 149 | 5,14 |
| 96 | Barnes BC, Cooper L, Kirkendall DT, McDermott TP, Jordan BD, Garrett WE Jr. Concussion history in elite male and female soccer players. Am J Sports Med. 1998 May-Jun;26(3):433-8. | 148 | 5,92 |
| 97 | Lehance C, Binet J, Bury T, Croisier JL. Muscular strength, functional performances and injury risk in professional and junior elite soccer players. Scand J Med Sci Sports. 2009 Apr;19(2):243-51. | 147 | 10,50 |
| 98 | Langberg H, Ellingsgaard H, Madsen T, Jansson J, Magnusson SP, Aagaard P, Kjaer M. Eccentric rehabilitation exercise increases peritendinous type I collagen synthesis in humans with Achilles tendinosis. Scand J Med Sci Sports. 2007 Feb;17(1):61-6. | 147 | 9,19 |
| 99 | Thorborg K, Krommes KK, Esteve E, Clausen MB, Bartels EM, Rathleff MS. Effect of specific exercise-based football injury prevention programmes on the overall injury rate in football: a systematic review and meta-analysis of the FIFA 11 and 11+ programmes. Br J Sports Med. 2017 Apr;51(7):562-571. | 146 | 24,33 |
| 100 | Boden BP, Kirkendall DT, Garrett WE Jr. Concussion incidence in elite college soccer players. Am J Sports Med. 1998 Mar-Apr;26(2):238-41. | 146 | 5,84 |
